# RABENOSYN separation-of-function mutations uncouple endosomal recycling from lysosomal degradation causing a distinct Mendelian Disorder

**DOI:** 10.1101/2021.10.03.21264281

**Authors:** Franziska Paul, Calista Ng, Shahriar Nafissi, Yalda Nilipoor, Ali Reza Tavasoli, Umar Bin Mohamad Sahari, Carine Bonnard, Pui-Mun Wong, Nasrinsadat Nabavizadeh, Mehrdad A. Estiar, Charles B. Majoie, Hane Lee, Stanley F. Nelson, Ziv Gan-Or, Guy A. Rouleau, Paul P. Van Veldhoven, Rami Massie, Raoul C. Hennekam, Ariana Kariminejad, Bruno Reversade

## Abstract

Rabenosyn (RBSN) is a conserved endosomal protein necessary for regulating internalized cargo. Here, we present genetic, cellular and biochemical evidence that two distinct *RBSN* missense variants are responsible for a novel Mendelian disorder consisting of progressive muscle weakness, facial dysmorphisms, ophthalmoplegia and intellectual disability. Using exome sequencing, we identified recessively-acting germline alleles p.Arg180Gly and p.Gly183Arg which are both situated in the FYVE domain of RBSN. We find that these variants abrogate binding to its cognate substrate PI3P and thus prevent its translocation to early endosomes. Although the endosomal recycling pathway was unaltered, mutant p.Gly183Arg patient fibroblasts exhibit accumulation of cargo tagged for lysosomal degradation. Our results suggest that these variants are separation-of-function alleles, which cause a delay in endosomal maturation without affecting cargo recycling. We conclude that distinct germline mutations in *RBSN* cause non-overlapping phenotypes with specific and discrete endolysosomal cellular defects.

## INTRODUCTION

Endocytosis allows cells to respond to their environment by actively transporting signalling molecules and integral cell surface proteins via invagination of the plasma membrane. This internalization process and the transport of endocytosed material to the right cellular destination is controlled at multiple levels (Cullen & Steinberg, 2018). Early endosomes (EEs) are typically defined as the compartment that first receives and sorts endocytosed cargo. Depending on the final destination, EE cargo is either packed into budding vesicles that recycle surface proteins and lipids back to the plasma membrane, or are fed into the endolysosomal pathway. In this pathway, EEs mature into late endosomes (LEs) to eventually fuse with lysosomes for targeted degradation (Scott et al, 2014; Gruenberg & Maxfield, 1995). Cargo may also be fed back to the plasma membrane or into the trans-Golgi network through budding vesicles from LEs. The continuous remodeling of endosomes can be defined by distinct events, including the switching from EE marker RAB5 to LE marker RAB7 (Chavrier et al, 1990; Deretic, 2005; Poteryaev et al, 2010; Rink et al, 2005; Woodman, 2000), the conversion of phosphoinositide (PI) from PI3P to PI(3,5)P2 (Poteryaev et al, 2010; Simonsen et al, 2001), and the fusion with endolysosomal vesicles (Scott et al, 2014; Gruenberg & Maxfield, 1995; Poteryaev et al, 2010; Simonsen et al, 2001; Mills et al, 1998).

Mendelian genetics has demonstrated that germline mutations in genes coding for components of the endolysosomal system play a significant role in neurodegenerative diseases (Schreij et al, 2015; Tsuji, 2010). For example loss-of-function variants in *GALC* cause Krabbe disease (MIM245200) (Pavuluri et al, 2017), highlighting the need for recycling of membrane building blocks through the endolysosomal pathway to prevent neurodegeneration (Spassieva & Bieberich, 2016). Likewise a loss of the endosomal protease Cathepsin D (CTSD, MIM610127), which is needed for intracellular protein breakdown in the lysosome, leads to congenital neuronal ceroid-lipofuscinosis, a neonatal lethal condition (Siintola et al, 2006).

Here we provide evidence that biallelic germline mutations in *RBSN* (MIM609511) are responsible for a new clinical entity. Rabenosyn (also known as ZFYVE20), is an evolutionarily-conserved multi-domain protein, which is ubiquitously expressed (Eathiraj et al, 2005). A whole body knockout is embryonic lethal in mice (MGI:4441653), *Drosophila* and *C. elegans* (Mottola et al, 2010; Gengyo-Ando et al, 2007). RBSN is instrumental for the endolysosomal degradation pathway (Nielsen et al, 2000; Navaroli et al, 2012) and is recruited to the EE surface through the binding of phosphoinositide substrates via its FYVE domain (Nielsen et al, 2000). Here, we present two novel homozygous *RBSN* alleles which specifically disrupt binding to its cognate substrate PI3P. Functional studies performed on patient cells revealed altered cellular localization of endogenous RBSN and delayed endosomal maturation. Our biochemical and cellular investigations suggest that these are separation-of-function alleles which delineate a novel clinical syndrome with features that are distinct from the complete RBSN knockout and hypomorphic alleles reported heretofore (Magoulas et al, 2018; Stockler et al, 2014).

## RESULTS AND DISCUSSION

### Two distinct recessive *RBSN* variants delineate novel Mendelian syndrome

We have previously reported three siblings with a unique familial phenotype born to consanguineous healthy Red. parents (Kariminejad et al, 2015). Prominent characteristics include multiple neurological phenotypes, mild developmental delay and characteristic facial traits (Figures 1A-B, Figure EV1A-B). Identity-By-Descent (IBD) mapping delineated a single homozygous block at 3p24.3-p25.3 spanning 57 genes, that was shared by all three siblings (Kariminejad et al, 2015). Here, we performed exome sequencing of the proband (II:1) and found two rare homozygous protein-coding variants within the IBD locus, that segregated with the disease (Kariminejad et al, 2015). While the variant p.Ser363Phe in *HACL1* was found to be none deleterious (see Expanded View), we examined a private single-nucleotide polymorphism (SNP) c.547G>A in *RBSN*, which leads to a replacement of a highly conserved Glycine with Arginine at position 183 in the FYVE domain of the RBSN protein (Figure 1C-D). Sanger sequencing confirmed complete segregation with the disease (Figure 1A). Residue Gly183 is invariant across all *RBSN* homologues in species from invertebrates to vertebrates and thus likely essential for *RBSN* function (Figure 1D). Pathogenicity prediction programs indicated the *RBSN* mutation as “deleterious” or “damaging”. This private SNP (rs376613564) was seen once at the heterozygous state in GnomAD (frequency < 1e^-5^), but did not occur homozygously in any large human cohort or in our in-house exome database.

**Figure 1.**
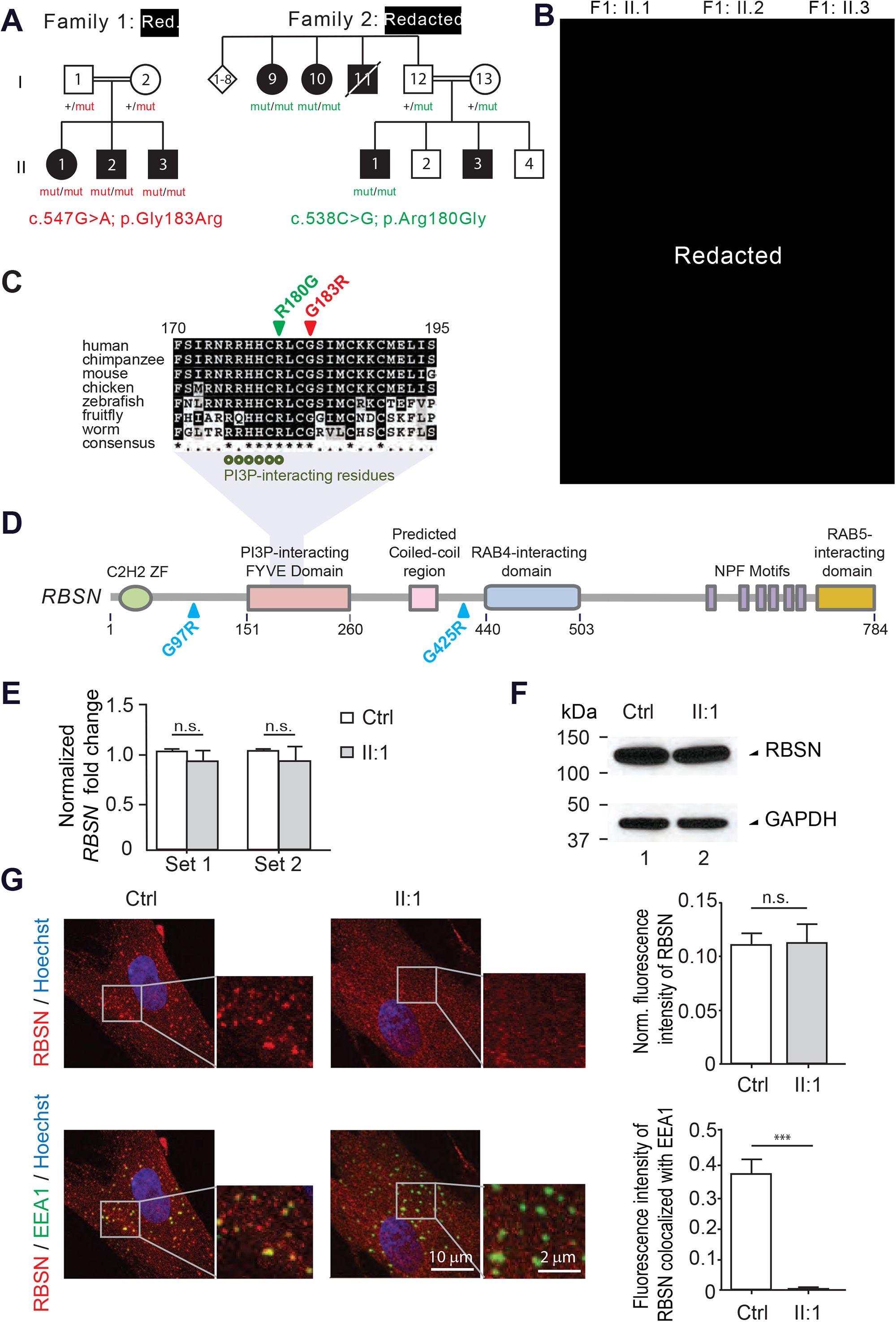
A Mendelian syndrome caused by RBSN^G183R^ or RBSN^R180G^ recessive variants affects RBSN localization to early endosomes. (A) Pedigrees of two unrelated consanguineous families from Red. and Red.. Affected individuals are marked in black. (B) Phenotypic characteristics of the affected Red. siblings with RBSN^G183R^ mutation. Prominent nasal bridge, ptosis, deeply set eyes, low hanging columella, small mouth, and slouching posture of II:1. See also Figure EV1. (C) Amino acid alignment of two highly conserved regions of RBSN. Green circles represent the residues directly interacting with PI3P based on EEA1-FYVE/PI3P structure (pdb: 1JOC). (D) Schematic diagram of RBSN protein showing the various functional domains. (E) No significant differences in *RBSN* transcript levels (normalized to *GAPDH)* were detected in patient fibroblasts relative to control cells. (Student *t*-tests, n=12) (F) Western blot against RBSN indicating no significant change of protein levels in patient fibroblasts relative to control cells. (G) Quantification of RBSN by immunofluorescence in patient derived fibroblasts. In patient fibroblasts RBSN (red) does not co-localize with the EE marker EEA1 (green). (Student *t*-tests: *** p<000.1, n=12 cells / 20 EES)

In an unrelated Redacted family of indigenous Cree descent with overlapping symptoms, we identified a homozygous variant c.538C>G by exome sequencing, that lies just nine nucleotides upstream of the mutation in the aforementioned Red. family (Figure 1A). This mutation causes a p.Arg180Gly substitution inside the highly conserved PI3P-binding domain (Figure 1D). This private c.538C>G variant was seen once in GnomAD (rs745678941, frequency < 1e^-5^) and recessively segregates with the disease in all available affected individuals (Figure 1A). Common phenotypes between the 6 affected individuals encompass developmental delay, intellectual disability, distal motor axonal neuropathy and facial dysmorphism; a comparative clinical table is provided for comparaison (Table EV1). The family did not consent to a skin biopsy or for pictures to be published; thus our experimental validation focuses on the allele from the Red. family.

Two previous studies have identified germline mutations in RBSN in children that had distinct clinical presentations. In 2014, Stockler and colleagues identified a homozygous missense RBSN p.Gly425Arg variant in a single child with developmental delay, epileptic encephalopathy, intellectual disability, microcephaly, dysostosis, intractable seizures and hematological and biochemical abnormalities (Stockler et al, 2014). In 2018, Magoulas and colleagues identified a homozygous missense p.Gly97Arg mutation leading to aberrant splicing of the *RBSN* transcript. With no RBSN protein observed in patient’s cells, this protein-null allele was recessively inherited in three siblings with severe intellectual disability, dysmorphic facial features and a syndromic form of congenital myelofibrosis with developmental defects (Magoulas et al, 2018). The presently reported mutations p.Arg180Gly and p.Gly183Arg are situated within the FYVE domain, which is responsible for substrate binding, while p.Gly425Arg and p.Gly97Arg lie in unannotated regions of RBSN (Figure 1D-E). The overlap in phenotype between these alleles is limited to nondescript traits such as disturbed growth, hypotonia, intellectual disability and facial dysmorphism (Table EV1). The degree of developmental delay and hypotonia varied considerably and other signs and symptoms were not congruent. We conjectured that *RBSN* mutations may lead to distinct diseases depending on their localization, as is often the case in large multidomain proteins (Worman & Bonne, 2007).

### Endogenous RBSN^G183R^ fails to localizes to early endosomes and cannot bind PI3P

When comparing primary fibroblasts from proband (II:1) to age- and ethnicity-matched control fibroblasts, no marked differences in *RBSN* transcript (Figure 1E) or protein levels (Figure 1F) were recorded, suggesting that RBSN protein stability is not affected by this private p.Gly183Arg mutation. However, immunofluorescence staining showed a disruption of the punctate staining for endogenous RBSN in the cytoplasm in RBSN^G183R^ cells (Figure 1G). Counterstaining with the EE marker EEA1 demonstrated that RBSN^WT^ is typically located in EEs, while RBSN^G183R^ failed to co-localize with EEA1. These results indicate that replacing Glycine with Arginine does not affect overall *RBSN* transcription or translation, but that the FYVE-specific variant specifically disrupts its localization to EEA1^+^ EEs. It should be noted here, that the EE compartment is heterogeneous and as such not all EEs, in particular recycling endosomes, contain EEA1 (Wilson et al, 2000; Navaroli et al, 2012).

RBSN belongs to a small family of FYVE domain-containing endosomal proteins (Nielsen et al, 2000). The FYVE zinc finger domain contains the highly conserved R+HHC+XCG motif, where “+” represents a charged residue and “X” represents any residue (Figure 1D-E). This motif specifically recognizes the phosphatidylinositol (PI) PI3P, which targets RBSN to early endocytic membranes (Nielsen et al, 2000; Gillooly et al, 2001). Since both RBSN^R180G^ and RBSN^G183R^ mutations fall within the FYVE consensus motif of RBSN, we investigated the effect of these mutations on the binding to PI3P.

Using the co-crystal structure of EEA1-FYVE/PI3P (pdb id: 1JOC) as a reference, we generated a homology model of RBSN-FYVE using the I-TASSER structure prediction software (Roy et al, 2010) (Figure 2A). The PI3P binding pocket consists of a beta hairpin formed by the C- and N-beta strands connected by a loop. Based on the reference structure (pdb id: 1JOC) Arginine 180 located on the N-strand of the beta hairpin is crucial for PI3P binding (Figure 2B). Amino acid Gly183 does not interact directly with PI3P but if forms a hydrogen bond with Cyst179 to maintain the structural integrity of the beta hairpin loop where half of the PI3P-interacting residues are found (Dumas et al, 2001; Gaullier et al, 2000). Changing a small uncharged Gly183 to a large positively-charged Arg, and vice versa changing Gly180 into Arginine, is likely to disrupt this loop conformation, causing the side chain of these otherwise invariant residues to turn away from PI3P or sterically hinder PI3P from engaging the FYVE domain.

**Figure 2.**
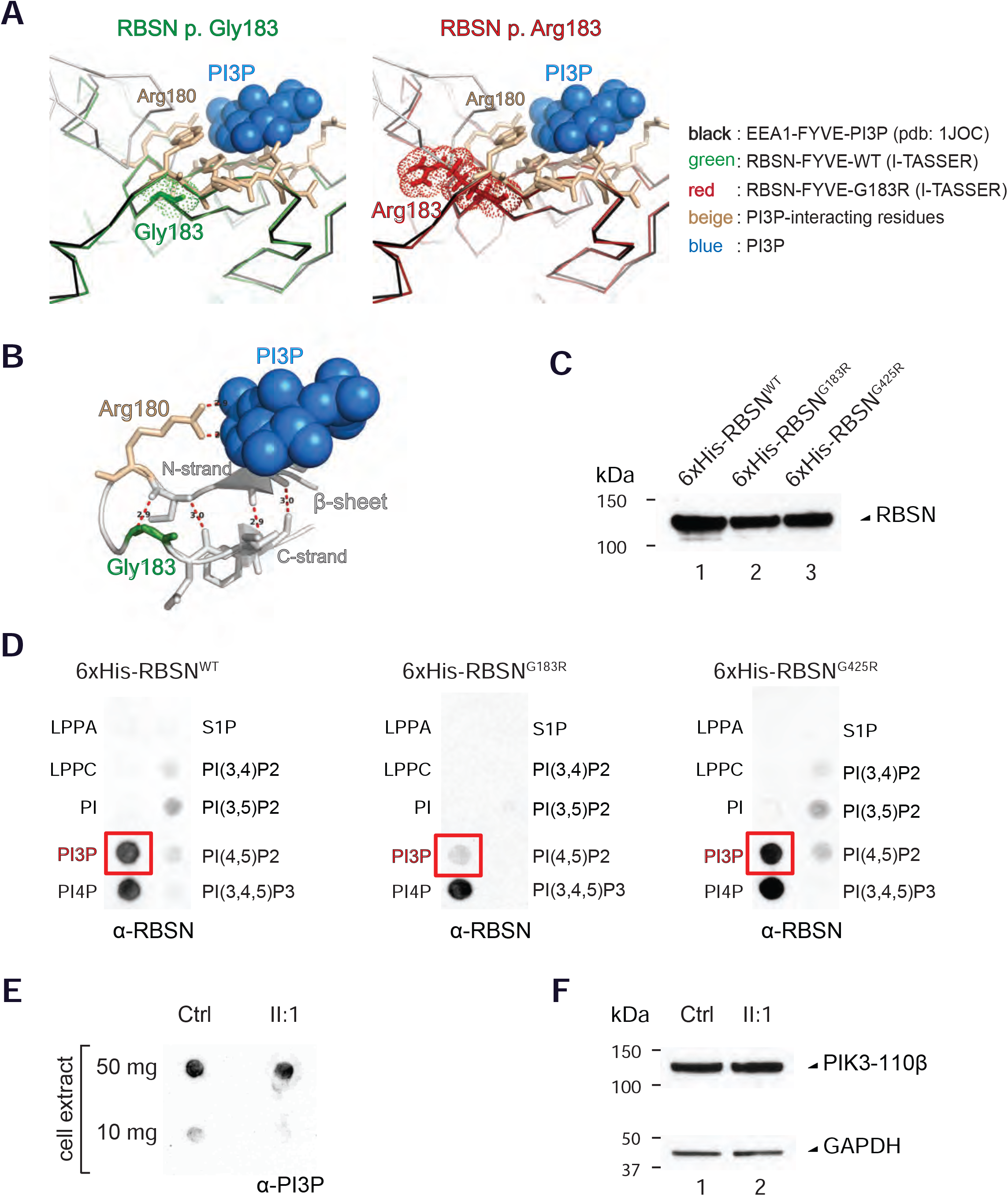
Mutant RBSN^G183R^ can no longer bind its natural substrate PI3P. (A) Superimposed structures of I-TASSER modeled RBSN (RBSN p.Gly183; green or RBSN p.Arg183; red) with the reference structure EEA1/PI3P (pdb: 1JOC; black). The amino acid 183 is adjacent to residues forming the PI3P binding pocket (pocket residues: beige sticks; partial PI3P: blue spheres) (B) Enlargement of the EEA1/PI3P (pdb: 1JOC) binding pocket highlights the importance of Gly183 (green) in maintaining the loop structure of the beta hairpin, allowing optimal binding of Arg180 (beige) with PI3P (blue). Red dotted lines show hydrogen bonding and distances in angstrom between residues. (C)Successful purification of recombinant 6xHis-RBSN proteins over a single-pass nickel Sepharose column. Western blot against RBSN. (D) *In vitro* PIP Strip™ binding assay using recombinantly purified RBSN proteins from C, demonstrating that the Gly183Arg mutation abrogates PI3P binding (red box). (E) Dot blot assay shows an equal amount of PI3P substrate in the cell extract of patient fibroblasts compared to control cells. (F) Western blot against Phosphoinositide 3-kinase (PIK3) generating PI3P substrate, shows equal amounts in patient fibroblasts and control cells.

To directly test this hypothesis, recombinant 6xHis-tagged-RBSN was overexpressed in HEK293T cells and affinity purified via a nickel-sepharose column (Figure 2C). Since the Arg180 is essential for PI3P binding, we examined the affinity of RBSN^G183R^ to that of another disease-causing mutation RBSN^G425R^ situated outside of the FYVE domain. Purified recombinant RBSN^G183R^ displayed dramatically reduced PI3P binding while recombinant RBSN^G425R^ bound with a similar affinity to that seen for RBSN^WT^ (Figure 2D). This defect is specific to the interaction with PI3P, as all three RBSN variants bound equally well to PI4P, which is not implicated in endosomal targeting (Nielsen et al, 2000).

To rule out that the absence of endogenous RBSN^G183R^ on EE is a result of insufficient PI3P in patient-derived fibroblasts, we performed a dot-blot assay. We detected comparable amounts of PI3P in lysates from both patient and control fibroblasts (Figure 2E). Moreover, the enzyme PI3-kinase (PI3K) responsible for the generation of PI3P molecules (Simonsen et al, 2001; Christoforidis et al, 1999) was equally expressed in both cell lines (Figure 2F). These biochemical results indicate that the absence of endogenous RBSN^G183R^ on EE may be the consequence of its loss of PI3P binding capacity, which we found to be specific to the p.Gly183Arg FYVE domain mutation and not the p.Gly425Arg variant. In support of our results is the finding that another point mutation in the FYVE domain of ZFYVE16, corresponding to p.Cyst163Ser of RBSN, also abrogated PI3P binding and disrupted its localization to EEs (Seet & Hong, 2001).

### Degradation, but not cargo recycling, of the endolysosomal pathway is impaired in RBSN^G183R^ cells

Endocytosed and processed cargo in early endosomes is either recycled back to the cell membrane or remains in the maturing endosome for degradation in lysosomes, both of which are dependent on RBSN (Navaroli et al, 2012). Here, we evaluated the impact of RBSN^G183R^ on both of these pathways with transferrin and dextran assays (Baravalle et al, 2005). To measure the rate of cargo recycling, patient (II:1) and control fibroblasts were pulsed with labeled transferrin. The rate of transferrin uptake and recycling were comparable between patient and control (Figure 3A). Thus we conclude that the endosomal recycling pathway is not affected in RBSN^G183R^ mutant cells. In line with this, accessory proteins involved in the endocytic pathway were expressed at comparable levels (Figure 3B). These findings contrast with previous reports, where a RBSN^G425R^ gain-of-function increased recycling speed (Stockler et al, 2014), and the RBSN^G97R^ loss-of-function slowed down endosomal recycling (Magoulas et al, 2018). This highlights a salient difference in the underlying molecular anomalies that underpin RBSN^R180G^ and RBSN^G183R^ specific phenotypes compared to those previously described. Presumably the localization of RBSN to EEA1^+^ EE is not determining for cell membrane targeted recycling, which speaks to the existence of heterogeneous EE sub-populations (Navaroli et al, 2012; Wilson et al, 2000).

**Figure 3.**
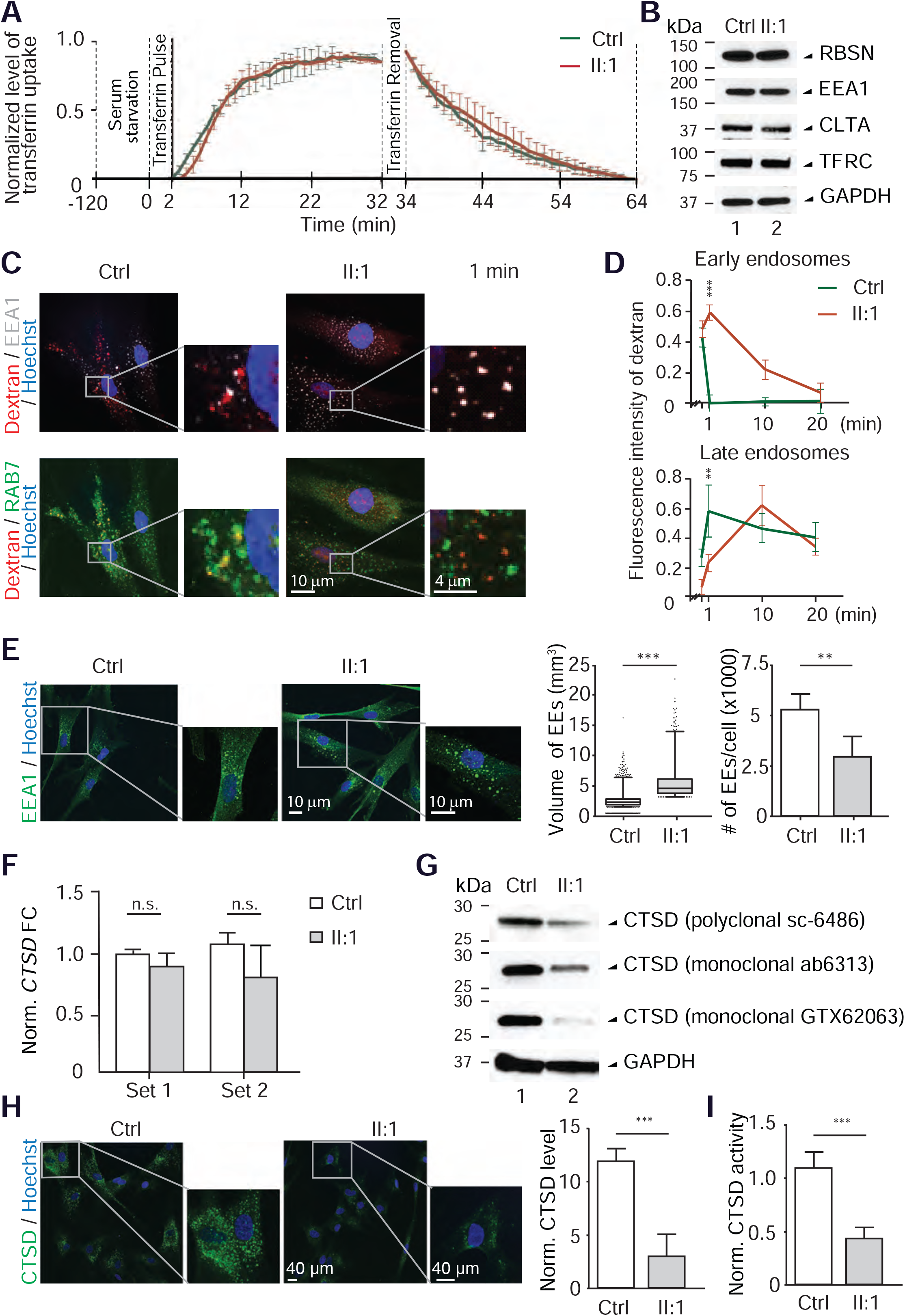
Loss of RBSN localization to early endosomes causes endosomal maturation delay and diminished Cathepsin D levels. (A)The rate of uptake and recycling of Alexa488-transferrin shows no significant differences between patient fibroblasts (red) and control cells (green) across time. (Two-way ANOVA, n=5). (B) Western blot of endosomal accessory proteins showing no significant differences between patient fibroblasts and control cells. (C) Immunofluorescence staining of control and patient fibroblasts 1-minute after pulsing with dextran. Dextran molecules were largely found in RAB7^+^ late endosome compartments in control cells (yellow), while in patient cells they remained in EEA1^+^ early endosomes (white). (D) Fluorescence intensity of dextran co-localized with early endosomes (top) and late endosomes (bottom) as a function of time (in minutes). Dextran transportation along the endolysosomal pathway was approximately 10-fold slower in patient cells compared to control cells. (Student *t*-test: ** p<0.001, *** p<000.1, n=5). (E) Immunofluorescence staining of patient fibroblasts showing increased size of individual EEA1-positive early endosomes (green) relative to those in control cells. The size (μm^3^) and number of EEs is quantified on the right. (Student *t*-tests: ** p<0.001, *** p<000.1, n=2396 EE). (F) QPCR detected no significant changes in *CTSD* transcript levels, normalized to *GAPDH*, in patient fibroblasts. (Student *t*-tests, n=12). (G) Western blot against CTSD shows a significant decrease in levels of the mature form of CTSD in patient fibroblasts. This was verified using three different commercially available antibodies. (H) Immunofluorescence staining confirms reduced levels of CTSD (green) in patient fibroblasts. Cellular CTSD levels are quantified on the right (p<0.0001, Student *t*-test). (I) CTSD enzymatic assay showed reduced levels of CTSD activity in patient fibroblasts as compared to control cells (normalized by input cell number, p<0.0001, Student *t*-test).

To assess whether the endosomal degradation pathway was impaired, we pulsed primary fibroblasts with dextran. After one minute, dextran molecules had travelled into the late endosome (LE) compartment (marked by RAB7) of control fibroblasts, while they remained in EEs (co-stained with EEA1) in RBSN^G183R^ patient cells (Figure 3C). In fact, patient cells exhibited a 10-minute delay to achieve a comparable dextran accumulation in LE (Figure 3D). This suggests that endosomes take longer to mature and hence cargo targeted for degradation may be transported less efficiently along the endolysosomal pathway in RBSN^G183R^ cells. In line with this, we found a significant enlargement of EE at the expense of their numbers in RBSN^G183R^ fibroblasts (Figure 3E). An increase of EE size has been previously reported after RBSN antibody-mediated neutralization or siRNA-mediated knockdown (Nielsen et al, 2000; Naslavsky et al, 2009), thus highlighting that the RBSN^G183R^ allele displays readily apparent loss-of-function vis-a-vis the endosomal degradation pathway.

Degradation, and thus clearance of protein aggregates, is carried out by endopeptidases in the lysosome. One essential endopeptidase, Cathepsin D (CTSD), is initially synthesized in the endoplasmic reticulum as an inactive glycosylated propeptide (Gieselmann et al, 1983). In the Golgi, pro-CTSD is then tagged with mannose-6-phosphate (M6P) which allows it to be sorted into the endolysosomal pathway (Pohlmann et al, 1995). The transport of pro-CTSD into an acidic environment in LEs and lysosomes finally induces proteolytic maturation of pro-CTSD into its active mature form (Zhila & Mary, 2014). While *CTSD* transcript levels remained comparable (Figure 3F), mature CTSD protein levels were markedly reduced in RBSN^G183R^ patient fibroblasts. This reduction of mature CTSD (∼28-32 kDa) was confirmed by three different commercial antibodies and by immunofluorescence (Figure 3G-H). In accordance with *RBSN* siRNA experiments (Naslavsky et al, 2009), RBSN^G183R^ likely fails to recycle M6P receptors from EEs to the trans-golgi network, thus inhibiting the release of immature CTSD into the endolysosomal pathway. Consistently, the total enzymatic activity of endogenous CTSD was more than halved in mutant RBSN^G183R^ cells relative to control fibroblasts (Figure 3I). Importantly, the RBSN^G425R^ allele also displayed a disruption of the endolysosomal degradation as documented by its poor CTSD activity (Stockler et al, 2014). These results suggest that RBSN^G183R^ leads to a partial loss of function in the endolysosomal degradation pathway which may be a likely driver of the disease.

While RBSN^G425R^ (Stockler et al, 2014), RBSN^G97R^ (Magoulas et al, 2018), RBSN^G183R^ and RBSN^R180G^, all cause Mendelian disorders characterized by developmental delay, intellectual disability, hypotonia, facial dysmorphism and delayed bone growth, they each present with additional symptoms that set them apart from one another (Table EV1). Combined, these domain-specific mutants unveil a decoupling of the endosomal recycling and endolysosomal degradation pathways in which RBSN functions. Thus discrete mutations in the same gene can bring about a gain-of-function (RBSN^G425R^), loss of function (RBSN^G97R^) or have no effect (RBSN^G183R^) on endosomal recycling. It is not uncommon for large multidomain proteins with multiple interacting partners to be responsible for several clinical entities. Mutations in *LMNA* (MIM150330), which are associated with over 10 different Mendelian disorders, exemplify this paradigm (Worman & Bonne, 2007). RBSN consists of six annotated protein domains. In addition to binding to RAB5, a master regulator of endosome biogenesis (Woodman, 2000; Zeigerer et al, 2012), endogenous RBSN was found to physically interact with 5 additional protein partners in 293T cells (Huttlin et al, 2015). We anticipate that the full spectrum of diseases associated with RBSN variants will become clearer as new alleles are delineated.

### FYVE-defective RBSN: A newly emerging lysosomal storage disorder?

Lysosomal Storage Disorders (LSD) encompass a family of diseases that arise from the inheritance of mutations affecting the homeostasis of the endolysosomal pathway. LSD can be grouped into two subtypes – primary and secondary (Platt et al, 2012). Primary LSD are caused by insufficiency of key lysosomal enzymes such as GALC in Krabbe disease (MIM245200) (Pavuluri et al, 2017). In contrast, secondary LSD have intact lysosomal enzyme functions but exhibit defective heterotypic organelle fusion resulting in the upstream accumulation of cargo. For example, in the secondary LSD Niemann-Pick disease type C2 (MIM607625), a mutation in *NPC2* leads to defective fusion of LEs with lysosomes, causing cellular accumulation of cholesterol (Goldman & Krise, 2009). Notwithstanding, the majority of LSD are caused by defects in the later part of the endolysosomal pathway (Platt et al, 2012), such that relatively few molecular and clinical insights have been gained from diseases affecting the early trafficking of cargo. The combination of CNS defects and progressive loss of psychomotor skills in the affected individuals homozygously carrying the p.Arg180Gly or p.Gly183Arg RBSN mutation, resemble clinical symptoms of LSD (Danon disease MIM300257, MLD MIM250100, Krabbe disease MIM245200). Consistent with this notion is our observation that mutant *RBSN*^*G183R*^ triggered a significant delay of EE maturation impacting the rate at which cargo, such as Dextran or CTSD, reached LE/lysosomes. Loss-of-function *RBSN* siRNA experiments have suggested that inactive CTSD gets trapped in the trans-golgi network (Naslavsky et al, 2009). The significant decrease in CTSD enzymatic activity observed in patient fibroblasts lends credence to this concept and warrants further investigation of CTSD levels in neuronal lineages where its activity has been shown to be most critical (MIM610127). Since neurons have extended cellular protrusions, they might be more reliant on RBSN for endosomal transport and long distance trafficking along axons. Henceforth we would like to suggest that this newly described syndrome may represent one of the first clinical entities that falls into the category of secondary LSD and whose molecular aetiology can be traced to early endosomal trafficking defects.

In conclusion, we provided clinical, genetic, cellular, molecular and biochemical evidence that a heretofore unknown Mendelian syndrome is likely the result of biallelic separation-of-function *RBSN* mutations specifically affecting its FYVE domain. This study helps uncouple the discrete functions of RBSN towards recycling of endocytosed cargo from its role in endolysosomal processes.

## MATERIALS AND METHODS

### Patients and clinical assessments

The clinical history and pertinent data on the evaluation of the affected siblings, including results of linkage studies, have been published elsewhere (Kariminejad et al, 2015). Genomic DNA samples were collected from peripheral lymphocytes from the siblings and their parents. A skin biopsy was obtained from affected individual II:1 of Family 1 (Figure 1A). Parents gave their informed consent for the execution of this study and the publishing of the results. This study has been ethically approved (A*STAR IRB #2019-087).

### Whole exome sequencing

Exome sequencing of patient II:1 was performed using the Illumina TruSeq Exome Enrichment Kit for exome capture with 1 μg of genomic DNA. Illumina HiSeq2000 platform with high-output mode was used as according to the manufacturer’s guidelines to obtain 100-bp paired-end reads at the UCLA Clinical Genomics Centre and at the UCLA Broad Stem Cell Research Center as previously described (Hu et al, 2014). After filtering, 60 homozygous, 132 compound heterozygous and 569 heterozygous variants were protein-changing with population minor allele frequencies of <1%. Of these, two rare homozygous variants were found in a shared IBD homozygous block at 3p24.3-p25.3 (Kariminejad et al, 2015).

### Mutation analysis

Direct Sanger sequencing of genomic DNA flanking the *RBSN* mutation was performed according to its sequence found in the GenBank and Ensembl databases. The primer sequences for *RBSN* (NM_022340) mutation screening are provided in Table EV1. Sanger sequence analysis was done with the BigDye Terminator cycle sequencing kit (Thermofisher Scientific, Waltham, MA), and products were run on a 3730 DNA Analyzer (Thermofisher Scientific).

### Cell culture

Fibroblast cells were generated from the patient skin biopsy. For expansion and maintenance, cells were cultivated in high glucose DMEM (Lonza, Basel, Switzerland) supplemented with 10% fetal bovine serum (FBS,) (Lonza), 1X penicillin/streptomycin (Gibco / Thermofisher Scientific) and 2 mM L-glutamine (Life Technologies / Thermofisher Scientific).

### Quantitative PCR

Total RNA was isolated from patient fibroblasts using the Trizol (Invitrogen) followed by RNeasy Mini Kit (Qiagen / Thermofisher Scientific). A total of 1 μg of RNA was reverse transcribed using Iscript™ cDNA Synthesis Kit (Bio-Rad, Hercules, CA), and transcript levels were determined using the ABI Prism 7900HT Fast Real-Time PCR System (ThermoFisher Scientific). Gene expression was normalized to the housekeeping gene *GAPDH*.

### Statistical analysis

Each experiment was repeated at least three times, and data were expressed as mean ± S.D. Differences among treatments were analysed by Student’s *t* test or two-way ANOVA test. Significant differences were considered those with a *p* value of ***< 0.0001, **< 0.001, *< 0.01.

### Western blot

Cells were lysed in RIPA extraction buffer supplemented with 1 mM dithiothreitol (DTT) and 1X Protease Inhibitor Cocktail (Roche, Basel, Switzerland). Protein concentrations of cleared lysates were measured using the BCA assay before an equal amount of protein was loaded on precast 4-20% Tris/Glycine/SDS polyacrylamide gradient gels (BioRad). Transferred PVDF membranes were blotted using antibodies against: 1:100 rabbit anti-HACL1 (GeneTex, Irvine, CA; GTX106858), 1:4000 mouse anti-GAPDH (Santa-Cruz Biotechnology, Dallas, TX; sc-47724), 1:250 polyclonal rabbit anti-RBSN (Sigma-Aldrich, St. Louis, MO; HPA044878), 1:100 rabbit anti-PI3K (Proteintech, Rosemont, IL; 20584-1-AP), 1:100 goat anti-EEA1 (Santa-Cruz; sc-6415), 1:55 rabbit anti-CLTA (Sigma-Aldrich; HPA050918), 1:200 anti-CTSD (GeneTtex; GTX62063, Abcam; ab6313, Santa-Cruz; sc-6486), 1 μg/ml mouse anti-TFRC (Life Technologies / Thermofisher Scientific; 13-6800). Secondary anti-mouse-HRP, anti-goat-HRP and anti-rabbit-HRP were used at 1:4000 before visualization on x-ray film with SuperSignal West Dura Chemiluminescent Substrate (ThermoFisher Scientific).

### Immunofluorescence detection

Primary cells were grown on a Nunc™ Lab-Tek™ 4-well glass chamber slide and fixed for 15 min in ice cold 4% (w/v) paraformaldehyde. Permeabilization using 0.6% (v/v) Triton-X in PBS was performed for 15 min, and then incubated with 1:55 rabbit anti-HACL1 (Abnova, Taipei, Taiwan; HPA055838), 1:67 rabbit anti-RBSN (Sigma-Aldrich; HPA044878), 1:50 goat anti-EEA1 (Santa-Cruz Biotechnology; sc-6415), 1 μg/ml mouse anti-RAB7 (Abcam, Cambridge, UK; ab50533); or 1:200 anti-CTSD (GenetTex; GTX62063) overnight at 4°C in 2% (w/v) BSA diluted with PBS. For visualization, 1:1000 secondary antibody conjugated to Alexa Fluor 568 or Alexa Fluor 488 (Molecular probes, Eugene, OR) was applied. Counter staining of nuclei was performed using 1X Hoechst. Images were captured using the MetaMorph™ software on the FV1000 Olympus confocal microscope equipped with a Leica camera and lens.

### Oxidation and esterification experiments

Fibroblasts were cultured in T25 flasks and incubated for 16-22 hr with 4 μM [1-^14^C]-radiolabeled fatty acids (specific activity between 6000 and 25,000 dpm/nmol), bound to BSA at a molar ratio 2:1 as described (Veldhoven et al, 1993). The synthesis of 2-methyl-[1-^14^C]-hexadecanoic acid, 3-methyl-[1-^14^C]-hexadecanoic acid, [1-^14^C]-lignoceric acid and 2-hydroxy-[1-^14^C]-octadecanoic acid has been described before (Veldhoven et al, 1993; Foulon et al, 2005). Radioactivity present in CO_2_ and the acid soluble material (or formate for α-oxidation substrates) produced in each flask was measured (Croes et al, 1996), as well as incorporation of radioactivity in phospholipids, triglycerides and cholesteryl esters.

### Protein expression and purification

Plasmid pCS2+ was used for expression of N-terminally tagged 6xHis-RBSN^WT^, 6xHis-RBSN^G183R^ and 6xHis-RBSN^G425R^. The exchange of amino acid at position 183 and 425 was performed using a QuikChange™ XL-II Mutagenesis kit (Agilent Technologies, Santa Clara, CA) as per manual instruction. Proteins were overexpressed in HEK293T by transfecting plasmids using Lipofectamine™ 3000 (Thermo Fisher Scientific) for 72 hr. Cells overexpressing GFP-tagged proteins were sent for immunofluorescence staining following the protocol described above. Cells expressing histidine-tagged proteins were purified on Nickel-Sepharose beads essentially following the manufacturer’s recommendation (GE Healthcare, Chicago, IL). Briefly, cell pellets were lysed in Lysis Buffer [50 mM Tris-HCl pH 7.4, 300 mM NaCl, 10 mM Imidazole, 1 mM EDTA, 1X Protease Inhibitor] and, after cell break, clarified lysates were applied to pre-equilibrated beads. Proteins were then eluted with Elution Buffer [50 mM Tris-HCl pH 7.4, 100 mM NaCl, 300 mM Imidazole, 1 mM EDTA, 1X Protease Inhibitor] after binding and washing.

### PIP-substrate binding assay

The assay was performed using PIP Strips™ essentially following the supplier recommendations (ThermoFisher Scientific). Briefly, purified 6xHis-RBSN proteins were allowed to bind PIP membrane pre-blocked with TBST in 3% fatty acid-free BSA (Sigma). Bound membranes were rinsed and then subjected to immunoblotting detection similar to those of western blot using 1:250 anti-RBSN (Sigma-Aldrich; HPA044878). BCA detection of both WT and mutant RBSN were performed to ensure an equal amount of proteins were present during binding.

### Dot blot assay

Cells were lysed in RIPA extraction buffer supplemented with 1 mM dithiothreitol (DTT) and 1X Protease Inhibitor Cocktail (Roche). Protein concentrations of cleared lysates were measured using the BCA assay and equal amounts of protein were spotted onto PVDF membrane. Blot was dried completely before probing with 5 ug/ml mouse anti-PI3P (Echelon Biosciences, Salt Lake City, UT; Z-P003). Secondary anti-mouse-HRP was used at 1:4000 before visualization on x-ray film with SuperSignal West Dura Chemiluminescent Substrate (Thermo Fisher Scientific).

### Transferrin uptake and recycling assay

Fibroblast cells were seeded overnight in a 35-mm petri-dish with 14-mm glass microwells (MatTek, Ashland, MA™) grown in complete media. Cells were starved in serum-free media for 2 hr before transferrin-Alexa488 (Thermo Fisher Scientific) was pulsed in. Live images were immediately recorded using a spinning disc confocal microscope (Leica, Wetzlar, Germany) for 30 min with 1 min intervals to record for the uptake of transferrin. Recycling of transferrin was performed by the complete removal of media containing transferrin by rinsing twice with PBS before serum-free media is added back. Recycling images were taken for the next 30 min duration with 1 min intervals.

### Endosome Maturation Assay

Cells were seeded in 12-well plates for overnight attachment, pulsed with 100 μg/ml of TexasRed Dextran of size 10,000 MW, rinsed with PBS once and then fixed with PFA to freeze the movement of dextran. Cells were then stained with markers (EEs: EEA1; LEs: RAB7) under the condition described earlier.

### CTSD enzymatic assay

Cellular CTSD activity was determined using a CTSD assay kit purchased from Sigma (CS0800). Cell extracts taken from an equal number of patient-derived and control fibroblasts were used. Assays were performed strictly as described in the manufacturer’s protocol.

## Data Availability

The data that support the findings of this study are available on request from the corresponding author B.R. The data are not publicly available due to restrictions to ensure privacy of research participants.

## ACKNOWLEDGEMENTS

We are indebted to the familes for their generous contribution to this study. We are grateful to our group members for discussion and advice. We are also thankful to Nithya Baburajendran for her insightful input on the structural models.

## AUTHOR CONTRIBUTIONS

FP, CN and BR conceived the project and wrote the paper. SN, YN, ART, CB, NN, MAE, CBM, RM and RCH diagnosed the patients and collected patient samples. FP, CN, UBMS, CB, PW, NN, MAE, HL, SFN and PPVV performed the experiments. ZG, AK and BR supervised the project.

## EXPANDED VIEW

While the main text focuses on the RBSN variant, we found a second protein-coding variant within the IBD locus through exome sequencing in the Red. family (Kariminejad et al, 2015). This variant was a c.1091C>T change in *HACL1*, encoding 2-hydroxyacyl-CoA lyase 1, an enzyme involved in peroxisomal alpha-oxidation (Veldhoven, 2010), changing Serine 363 into phenylalanine. Peroxisomal tests including measurements of C22-C24 ratio, C22-C26 ratio and pristanic and phytanic acid levels, yielded normal results in all affected patients (Kariminejad et al, 2015). In cultured patient fibroblasts, degradation of 3-methyl branched and 2-hydroxy long chain fatty acids proceeded normally (Figure EV1C-D), indicating that apha-oxidation was not affected. Pathogenicity prediction programs such as M-CAP (Jagadeesh et al, 2016), SIFT (Ng & Henikoff, 2003), and POLYPHEN-2 (Adzhubei et al, 2010) predicted the *HACL1* mutation to be “tolerable” or “non-damaging” to protein function. Expression and localization of endogenous HACL1 in primary fibroblasts from proband II:1 were similar to those in age- and ethnicity-matched control cells (Figure EV1E-G). We also note that knocking out *HACL1* in mice (MGI:1929657) leads to cardiovascular defects which do not resemble the clinical phenotype observed in this family. Thus we concluded that the c.1091C>T *HACL1* variant was unlikely to be pathogenic.

## EXPANDED VIEW FIGURE LEGENDS

**Figure EV1.**
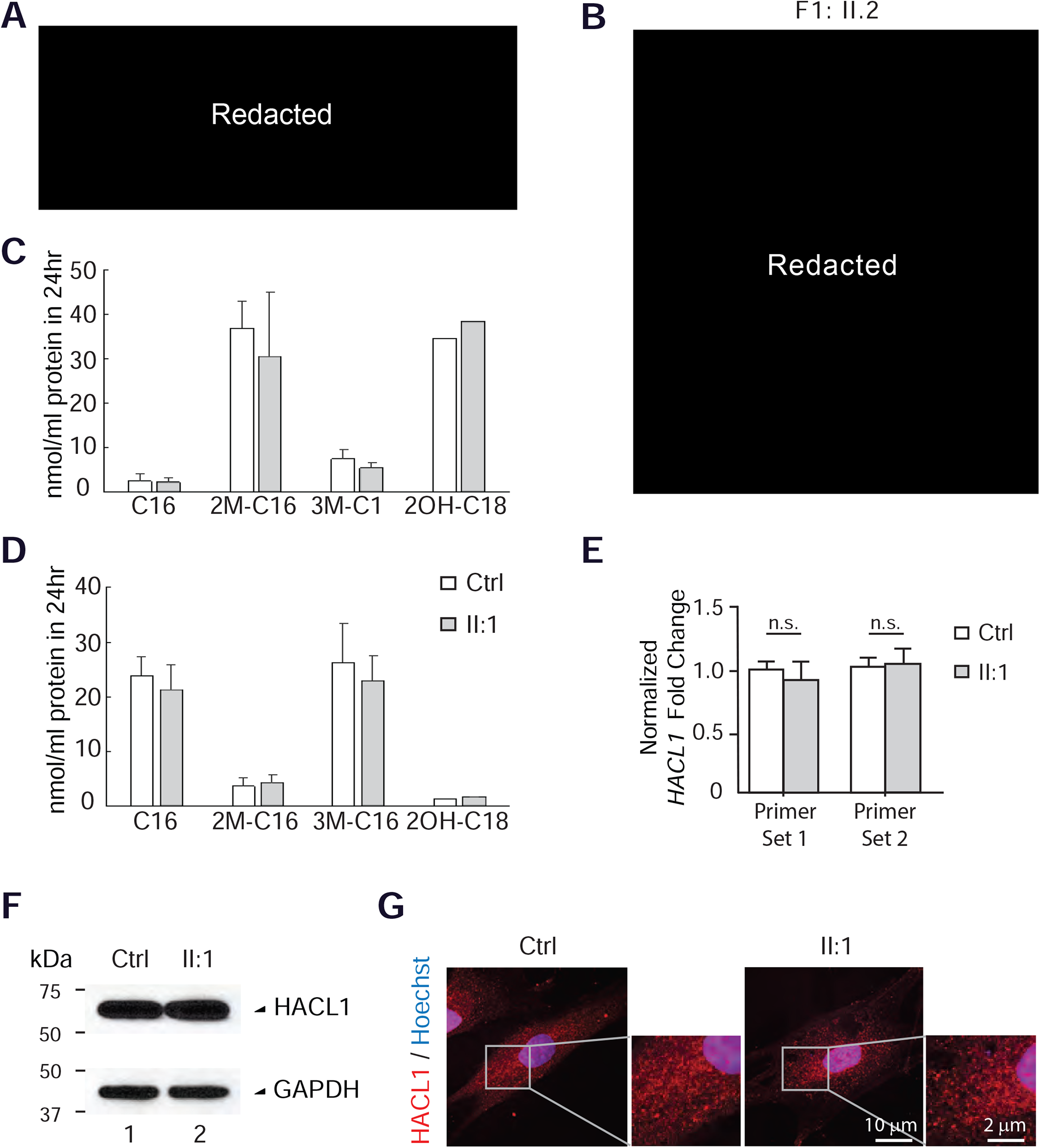
HACL1 variant does not show any pathogenic consequences. (A)Facial features of the three siblings at a young age include highly arched eyebrows and prominent nasal bridge. (B)Proband II:2 is lacking outermost incisors, has a high arched palate, shows overgrowth of the second toe over the first one and has a fused simian crease on the palm of his hand. (C)Formation of radioactive degradation products by fibroblasts incubated with 1-14C-labeled hexadecanoic acid (C16, mitochondrial beta-oxidation); 2-methylhexadecanoic acid (2M-C16, peroxisomal beta-oxidation); 3-methylhexadecanoic acid (3M-C16, peroxisomal alpha-oxidation); 2-hydroxyoctadecanoic acid (2OH-C18, peroxisomal alpha-oxidation) and lignoceric acid (peroxisomal beta-oxidation; data not shown). Data, normalized to protein content (100 - 200 μg/T25 flask), are derived from 2 (control) and 4 (patient) T25 flasks, incubated on two different days, except for 2OH-C18 (1 control, 2 patient flasks). (D)Incorporation of label into lipids (sum of cholesteryl esters, triglycerides and phospholipids) by fibroblasts incubated with 1-14C-labeled fatty acids described above. Data, normalized to protein content (100 - 200 μg/T25 flask), are derived from the incubations described in panel E (E)QPCR detects no significant differences in *HACL1* transcript levels, normalized to *GAPDH*, in patient fibroblasts relative to control cells. (Student *t*-tests, n=12). (F)Western blot against HACL1 shows no significant change of protein levels in patient fibroblasts relative to control cells. (G)Immunofluorescence staining of patient fibroblasts showed no significant difference in the cellular localization of HACL1 punctate stainings (red) relative to control cells.

**Table EV1.** Comparison of patient phenotypes between two diseases caused by distinct *RBSN* mutations. Symbols: +: present; -: not present; NE: not examined. Fractions indicate in how many patients the symptom was present.

|  | Family 1 | Family 2 | Stockler *et al.*, (2014) | Magoulas *et al.*, (2018) |
| --- | --- | --- | --- | --- |
| **Gene** | *RBSN* (MIM609511) |  |  |  |
| Inheritance | Homozygous | Homozygous | Homozygous | Homozygous |
| Mutation at cDNA level (NM_022340.3) | c.547G>A | c.538C>G | c.1273G>A | c.289G>C |
| Mutation at protein level (NP_001289307.1) | p.Gly183Arg | p.Arg180Gly | p.Gly425Arg | protein-null (exon 5 skipped) |
| **Overlapping symptoms** |  |  |  |  |
| Developmental delay | + | 2/4 | + | + |
| Intellectual disability | + | 3/4 | +++ | +++ |
| Inability to talk | - | - | + | + |
| Seizure/Tremor | 1/3 | - | Controlled with diet & medication | NE |
| Hypotonia/muscle weakness | + | + | +++ | +++ |
| Inability to sit | - | - | - | + |
| Inability to stand | - | - | + | + |
| Inability to walk | - | - | + | + |
| Slouching posture | + | - | NE | NE |
| Spastic paraparesis | NE | 2/4 | NE | NE |
| Facial dysmorphism | + | 2/4 | + | + |
| Delayed bone growth | + | - | + | 2/3 |
| **Non-overlapping symptoms** |  |  |  |  |
| Delayed myelination | + | 1/4 | - | 1/3 |
| Hippocampal atrophy | 1/3 | - | - | NE |
| Leukoencephalopathy | + | - | - | NE |
| EMG: myopathic pattern | + | - | - | NE |
| EMG: motor sensory neuropathy | NE | + | NE | NE |
| Ophthalmoplegia | + | 1/4 | - | NE |
| High lactate dehydrogenase | + | 1/4 | - | NE |
| High alkaline phosphatase | + | - | - | NE |
| High creatinine phosphokinase | + | 1/4 | - | NE |
| Elevated Vitamin E | + | NE | - | NE |
| Microcephaly | - | 1/4 | + | NE |
| Enlargement of 3<sup>rd</sup> ventricle | - | 2/4 | + | NE |
| Enlargement of lateral ventricles | NE | + | NE | NE |
| Acute neutropenia | - | - | + | + |
| Foamy macrophages | - | - | + | - |
| Reticulin fibrosis | - | NE | + | + |
| Vitamin B12 deficiency | - | - | + | NE |
| Hyperlipidemia | - | 1/4 | + | + |
| Microalbuminuria | - | NE | + | NE |
| Osteopenia | NE | NE | + | 2/3 |
| Anemia | - | - | - | + |
| Thrombocytopenia | - | - | - | 2/3 |
| Hepatomegaly | - | - | NE | + |
| Extramedullary hematopoiesis | NE | - | NE | +++ |
| Microphthalmia | - | - | NE | + |
| Optic nerve hypoplasia | NE | - | NE | + |
| Poor visual response | 1/3 | - | NE | + |
| Thin corpus callosum | NE | - | NE | + |
| Myelofibrosis | NE | - | - | +++ |
| XY Sex reversal | NE | NE | NE | + |
| Cerebral atrophy | NE | - | NE | + |
| Tracheomalacia | NE | - | NE | 2/3 |
| Gastrostomy feeding dependent | - | - | - | + |
| Hearing impairment | NE | - | NE | 1/3 |
| Vocal cord paralysis | NE | 1/4 | NE | NE |

**Table EV2.**
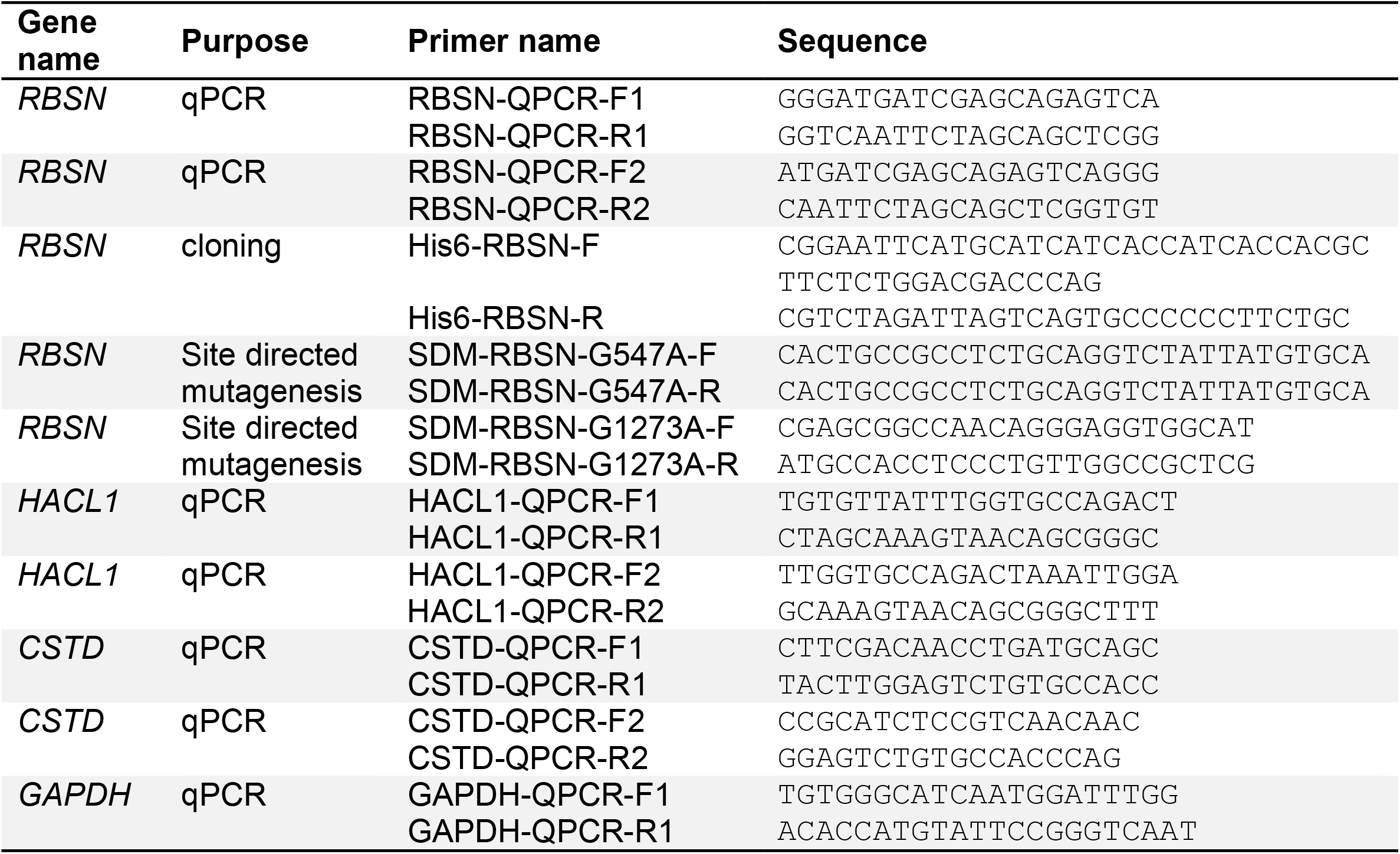
Primers used in this study

